# Risk Score Models for Unplanned Urinary Tract Infection Hospitalization

**DOI:** 10.1101/2023.08.06.23293723

**Authors:** Nasrin Alizadeh, Kimia Vahdat, Sara Shashaani, Julie L. Swann, Osman Ozaltin

**Affiliations:** Edward P. Fitts Department of Industrial and Systems Engineering, North Carolina State University, Raleigh, United States

**Keywords:** Risk score model, Urinary tract infection, Centers for Medicare and Medicaid Services

## Abstract

Annually, urinary tract infections (UTIs) affect over a hundred million people worldwide. Early detection of high-risk individuals can help prevent hospitalization for UTIs, which imposes significant economic and social burden on patients and caregivers. We present two methods to generate risk score models for unplanned UTI hospitalization. We utilize a sample of patients from the insurance claims data provided by the Centers for Medicare and Medicaid Services to develop and validate the proposed methods. Our dataset encompasses a wide range of features, such as demographics, medical history, and healthcare utilization of the patients along with provider quality metrics and community-based metrics. The proposed methods scale and round the coefficients of an underlying logistic regression model to create scoring tables. We present computational experiments to evaluate the prediction performance of both models. We also discuss different features of these models with respect to their impact on interpretability. Our findings emphasize the effectiveness of risk score models as practical tools for identifying high-risk patients and provide a quantitative assessment of the significance of various risk factors in unplanned UTI hospitalizations such as admission to ICU in the last 3 months, cognitive disorders and low inpatient, outpatient and carrier costs in the last 6 months.

## Introduction

Urinary tract infections (UTIs) affect around 150 million people worldwide each year, including 11 million in the United States (US) [1,2]. A study in the US estimated that UTIs were associated with over 10.5 million physician visits and 2-3 million emergency department visits in 2007 [3]. Furthermore, UTIs account for a substantial number of antibiotic prescriptions [4,5]. Effective and timely outpatient care can help reduce the likelihood of hospitalization for UTI, as it is considered an ambulatory care sensitive condition [6]. Hospitalizations are significantly more expensive than outpatient or primary care, thus potentially preventable hospitalizations are closely monitored to evaluate the efficiency of health systems [7]. UTIs caused about 380,600 potentially preventable adult inpatient stays, costing 2.55 billion dollars in the US in 2017 [8].

Women are more likely to develop UTIs than men, with an estimated 50-60% of women experiencing at least one UTI in their lifetime [9]. Women are also more likely to experience recurrent UTIs, which can further increase the risk of hospitalization [10]. In addition to sex, other risk factors for UTIs include age, urinary catheterization, urinary tract abnormalities, pregnancy, and history of UTIs [3,11,12]. Adults with cognitive impairment [13] and individuals with conditions such as chronic kidney disease, diabetes, and immunosuppression are also at increased risk for UTIs [3,14]. UTIs are also prevalent after a kidney transplant with two main triggers being vesicoureteral reflux and the use of immunosuppressive drugs [14]. These population groups may require intensive management of UTIs and closer monitoring to prevent hospitalization.

Predictive analysis of clinical and healthcare utilization data can effectively reduce healthcare costs and improve the quality of care for UTIs. Taylor et al. [15] developed machine learning models for predicting UTI among patients in the emergency department (ED) using demographic information, vitals, laboratory results, medications, past medical history, chief complaint, and structured historical and physical exam findings. Their best performing method achieves 0.904 Area under the ROC Curve (AUC), and 87.5% accuracy, however, it is limited to the patients in the emergency department with urine culture results. Other UTI-related data-driven models include predicting the risk of acquiring UTI in hospitalized patients [16], and predicting drug effectiveness in treating UTI [17,18]. Mao et al. [19] proposed a hierarchical clustering approach for predicting unplanned hospitalizations for UTI using Medicare fee-for-service claims data.

The interpretability of data-driven healthcare decision aid tools is critical for troubleshooting and understanding the model results [20]. However, maintaining high prediction performance while improving interpretability is challenging. Several strategies are proposed to build machine learning models that are easily understandable and usable by healthcare providers and policymakers [21,22]. In this study, we develop two risk score models to predict unplanned hospitalizations for UTI using claims data from Centers for Medicare and Medicaid Services (CMS) on healthcare utilization, including hospitalizations and physician visits. We augment this administrative claims data by community-level variables and provider quality metrics obtained from publicly available data sources. Risk score models provide an easy-to-use and practical risk assessment tool where integer points assigned to model features are summed together [20], and the total score is used to make predictions. Such models are widely used in healthcare [23–25], finance [26], and criminal justice [27]. We define unplanned hospitalizations based on the Prevention Quality Indicator (PQI) criteria of the Agency for Healthcare Research and Quality (AHRQ) [28]. We present computational experiments to evaluate the prediction performance of the proposed models. We also discuss different features of these models with respect to their impact on interpretability. Our findings emphasize the effectiveness of risk score models as practical tools for identifying high-risk patients and provide a quantitative assessment of the significance of various risk factors in unplanned UTI hospitalizations.

### Problem

Unplanned hospitalizations for urinary tract infections impose significant economic and social burden on patients and caregivers.

### What is Already Known

Although predictive analysis of clinical data has been proposed to improve the quality of care for UTIs in prior studies, a prominent limitation of these explorations is that they only consider emergency department or hospitalized patients. Furthermore, they don’t emphasize interpretability, which is critical for troubleshooting and understanding the model results of data-driven healthcare decision aid tools.

### What This Paper Adds

In this study, we develop two risk score models to predict unplanned hospitalizations for UTI using claims data from Centers for Medicare and Medicaid Services on healthcare utilization, including hospitalizations and physician visits. We augment this administrative claims data by community-level variables and provider quality metrics obtained from publicly available data sources. Risk score models provide an easy-to-use and practical risk assessment tool where integer points assigned to model features are summed together, and the total score is used to make predictions.

## Materials and Methods

### Data

This is a retrospective study using Medicare Limited Data Sets (LDS) from 2008 to 2012. CMS made this fully anonymized data available to our group on 1/16/2020 for an Artificial Intelligence Health Outcomes Challenge to predict unplanned hospital and skilled nursing facility admissions and adverse events based on Medicare Fee-for-Service (FFS) Parts A and B administrative claims [29]. The data set contains records from 2008–2012 for a random 5% sample of Medicare beneficiaries. We don’t have access to information that could identify individual participants during or after data collection. The overall project with human subjects research was reviewed by the Institutional Research Board at North Carolina State University (IRB Protocol 20528). Data storage, maintenance, and protection are governed by the Data Use Agreement between the university and the CMS.

We exclude beneficiaries without any inpatient or Skilled Nursing Facility (SNF) claim or who live in a nursing home. We also exclude beneficiaries who live in areas other than the 53 US states and territories. Finally, we exclude beneficiaries with urinary cancer, end-stage renal disease (ESRD) related claims, less than 2,000 USD total annual cost in carrier, inpatient, outpatient, and SNF claims or cost abnormalities including those with carrier claims but zero cost in carrier file, and outpatient claims but zero cost in carrier or outpatient files.

We do not consider the first 3 months and the last month of 2011 and 2012 in our analysis to ensure consistent computation of rolling horizon variables, e.g., inpatient cost in the last 3 months. We also filter records of patients who are dead, in hospital or hospice at the beginning of the current (observation) month, or who are enrolled in managed care at the beginning of the current month. We use data from April to November of 2011 for training, and data from 2012 during the same period for testing and performance evaluation.

In addition to the medical claims, our data set contains relevant variables from publicly available datasets such as Population Census Elderly Living [30]; Immunization [31]; County Health Rankings [32]; Hospital and Nursing Home Compare Dataset [33,34]; Weekly U.S. Influenza Surveillance Reports [35]. We also computed 285 Clinical Classification System (CCS) variables based on the ICD-9 diagnosis codes identified by AHRQ and added to our data. An extended description of our data can be found in [19]. The final data used for the analysis contains 821 variables including demographic characteristics and medical history of beneficiaries, their healthcare utilization in the past 1-6 months, as well as provider quality metrics, census information, and community-based regional public health metrics (e.g., flu vaccine coverage in an area); (see Table 1).

**Table 1.**
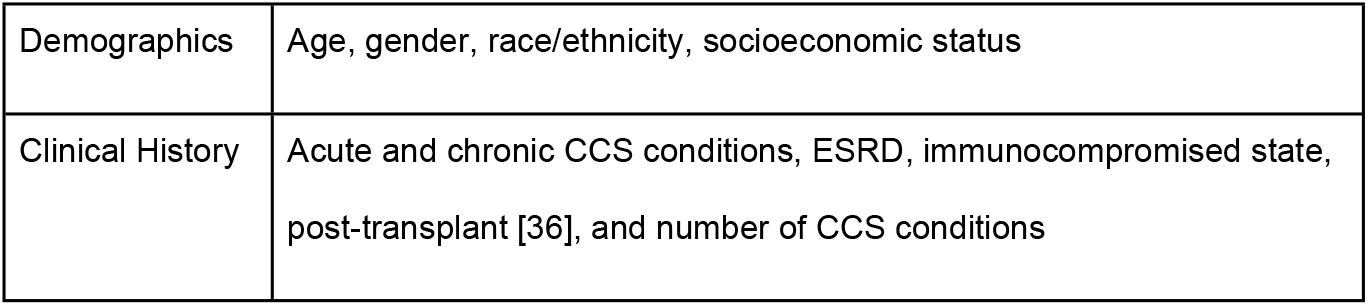
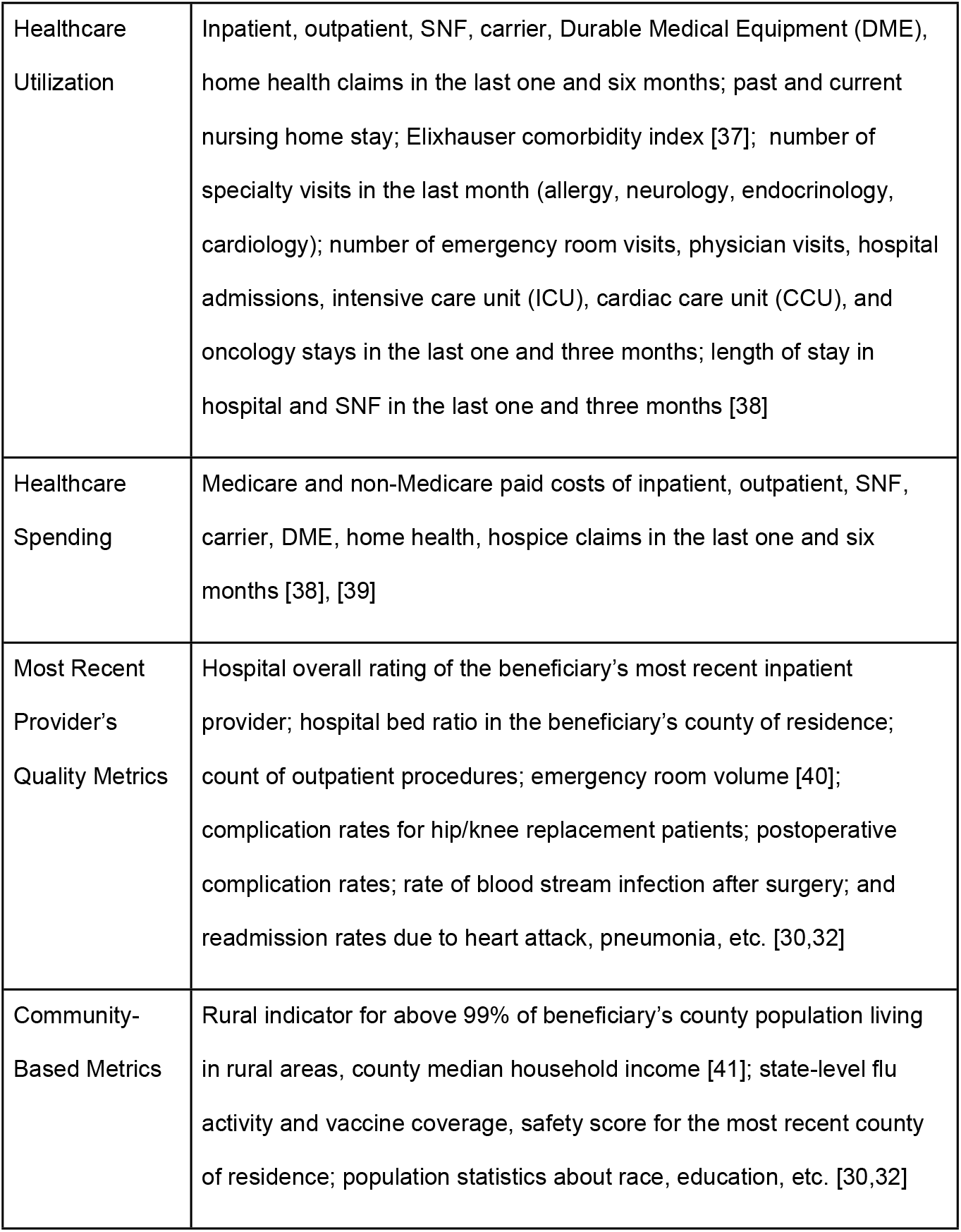
Summary of the variables considered in the model. Socioeconomic status indicates whether a beneficiary has supplemental insurance.

|  |  |
| --- | --- |
| Demographics | Age, gender, race/ethnicity, socioeconomic status |
| Clinical History | Acute and chronic CCS conditions, ESRD, immunocompromised state, post-transplant [36], and number of CCS conditions |
| Healthcare Utilization | Inpatient, outpatient, SNF, carrier, Durable Medical Equipment (DME), home health claims in the last one and six months; past and current nursing home stay; Elixhauser comorbidity index [37]; number of specialty visits in the last month (allergy, neurology, endocrinology, cardiology); number of emergency room visits, physician visits, hospital admissions, intensive care unit (ICU), cardiac care unit (CCU), and oncology stays in the last one and three months; length of stay in hospital and SNF in the last one and three months [38] |
| Healthcare Spending | Medicare and non-Medicare paid costs of inpatient, outpatient, SNF, carrier, DME, home health, hospice claims in the last one and six months [38], [39] |
| Most Recent Provider's Quality Metrics | Hospital overall rating of the beneficiary's most recent inpatient provider; hospital bed ratio in the beneficiary's county of residence; count of outpatient procedures; emergency room volume [40]; complication rates for hip/knee replacement patients; postoperative complication rates; rate of blood stream infection after surgery; and readmission rates due to heart attack, pneumonia, etc. [30,32] |
| Community-Based Metrics | Rural indicator for above 99% of beneficiary's county population living in rural areas, county median household income [41]; state-level flu activity and vaccine coverage, safety score for the most recent county of residence; population statistics about race, education, etc. [30,32] |

#### Observation generation

Each observation (row) corresponds to a patient-month. The number of observations for a patient varies between 1 and 8 in 2011 and 2012. To ensure that the model only employs historical data to predict future events, the response for each patient-month is the unplanned UTI hospitalization in the next month. We identify unplanned UTI hospitalizations based on the PQI 12 criteria (urinary tract infection admission rate) defined by AHRQ [28].

We consider beneficiaries who had at least one claim with UTI diagnosis based on the CCS variables since 2008. This group of patients has a 46.45% coverage of all events (unplanned UTI hospitalizations) in the data with 0.71% prevalence rate. Population statistics of these patients are presented in Table 2.

**Table 2.** Descriptive statistics of the beneficiaries with a history of UTI considered in this stud*y*.

|  | 2011 | 2012 |
| --- | --- | --- |
| Total beneficiaries | 64,673 | 60,574 |
| Gender (%) |  |  |
| Male | 23.1% | 23.2% |
| Female | 76.9% | 76.8% |
| Age categories (%) |  |  |
| < 65 | 0.8% | 0.1% |
| 65 - 69 | 11.3% | 11.6% |
| 70 - 74 | 14.6% | 14.9% |
| 75 - 79 | 17.3% | 17.6% |
| 80 - 84 | 21.2% | 20.5% |
| >84 | 34.8% | 35.3% |
| Race and Ethnicity (%) |  |  |
| Unknown | 0.2% | 0.2% |
| White | 85.5% | 85.4% |
| Black | 9.4% | 9.4% |
| Other | 1% | 1% |
| Asian | 1.2% | 1.3% |
| Hispanic | 2.1% | 2.2% |
| North American Native | 0.6% | 0.5% |
| Comorbid Conditions (%) |  |  |
| Diabetes Mellitus | 42.3% | 44.8% |
| Delirium and Cognitive Disorders | 56.2% | 59.7% |
| Heart disease | 93.6% | 94.5% |

## Methods

Different approaches can be used for assigning points to model features when developing a risk score. In this study, we apply two methods. The first method, referred to as *integerized logistic regression (LR)*, aims to develop a risk score with a given total score range. The second method, referred to as the *credit scorecard model*, regulates the increment in the total score associated with a specific level of increase in the odds of the outcome.

In the integerized LR method, variables are transformed into binary variables (e.g., six binary variables for each category of age), an LR model is built, and its coefficients are scaled and rounded. Treating each category within a variable as a dummy binary variable results in high collinearity between variables. We address this issue by limiting the number of features to 10 using a LASSO penalty in training [42]. In the credit scorecard model, each categorical value of a variable is replaced by its weight of evidence (WOE) [43]. We then select 15 variables with the highest information value (IV). The IV of a variable shows its strength as a predictor and is calculated as the weighted sum of the WOE values for all categories of that variable. The weight of each category is determined based on the difference between its frequency among events and non-events [44]. We use the logistic regression LASSO method to select a maximum of 10 variables from 15 variables with the highest IV. The coefficients of this model are then scaled and rounded to enable risk score calculation with integer values. Unlike the integerized LR method, the credit scorecard model assigns a score to each category of a variable included in the model allowing users to evaluate how being in each category affects the overall score. Each method is explained in further detail below after describing the variable categorization.

### Variable Categorization

Our data consists of binary, continuous, and categorical variables. We discretize continuous variables because the credit scorecard model requires categorical inputs. Furthermore, categorization improves the interpretation of the cost and the number of claim variables whose values are concentrated around zero with only a few large values. We use a binning algorithm to create categories with monotone WOE. For each category of a variable, WOE is calculated as the log of the ratio of its frequency among events to that among non-events. In our case, the event refers to an unplanned UTI hospitalization. Thus, a higher WOE for a category implies a higher risk for unplanned UTI hospitalization.

### Integerized LR Method

Let *X*_1_, *X*_2_,…, *X*_*d*_ denote vectors of the considered variables for each beneficiary, and *Y* be the binary response variable indicating the unplanned UTI hospitalization. We assume *Y* follows a Binomial distribution, which takes the value of one with probability *p* = *Pr*(*Y* = 1|*X*_1_, *X*_2_, …, *X*_*d*_), and define the general logistic regression model as

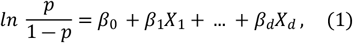

where *β*_0_ is the intercept and *β*_1_,…, *β*_*d*_ are parameters corresponding to each independent variable. These parameters can have fractional values. In the proposed approach, we scale and round them to integers as below.

Define *c* = |*β*_0_|⍰ *a* as a scaling factor, where *a* is a positive constant. Scaling the right-hand side of (1) by *c* yields,

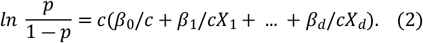

We round the scaled parameters in the parentheses to the closest integer value. Hence, the score of each feature is *β*′_*k*_ = [*β*_*k*_/*c*] for *k* = 0,1, …, *d*, and the approximated log of odds is given by

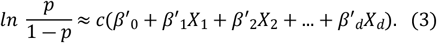

We refer to (3) as integerized LR model. The risk score for patient *i* is given by

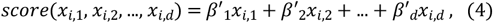

where features are generated by categorizing the original covariates as discussed before. Thus, *x*_*i*,1_, *x*_*i*,2_, …, *x*_*i,d*_ are binary variables. Subsequently, the conversion of a risk score to probability is through the logit function, i.e.,

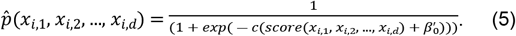

Using the proposed method, it is possible to achieve a specific range of total scores. Denote the min and max total score by *S*_*min*_and *S*_*max*_, respectively. We choose parameter *a* such that the total score range *R* = *S*_*max*_ ― *S*_*min*_ is larger than a given value *R*^∗^. Specifically, we use the following steps:

i. Initialize *a* such that

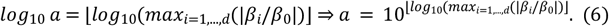

Here *a* is defined with base 10 logarithms to capture the order of magnitude difference between the maximum estimated coefficient and the intercept. The floor operator ensures that *a* is a power of 10 for simplicity of rounding.
ii. While *R* is smaller than *R*^∗^ set *a* = *a*/2. Step (i) sets the value of *a* as the order of magnitude that the largest coefficient is larger than the intercept of the logistic regression model. For example, if the intercept is 100 times smaller than the largest coefficient, then *a* = 100, but if they are in the same order of magnitude, then *a* = 1. The reason behind this choice is that we want to force the variables selected for the risk score model to have a significant contribution in changing the risk of hospitalization. The coefficients of those features that only marginally change the risk, will hence be zeroed out. Step (ii) decreases the value of *a*, hence the scaling factor c, to allow smaller coefficients in the model in order to achieve the target minimum total score range. In the computational analysis, the scaling factor c is set to 0.83 based on the results of the LR model.

### Credit Scorecard Model

The credit scorecard model was first developed to assess the risk of defaulting on a debt in the finance literature [43]. We begin by applying a data transformation that replaces each categorical value with its WOE. We then select a subset of the variables with the highest IV. After these two steps, a LASSO logistic regression model is trained allowing for a maximum of *d* variables. The total score in the scorecard model is a linear function of the log of odds, that is *S* = *A* + *B* × *log*(*odds*). Using the LR model with WOEs as variable values, we can expand this formula as:

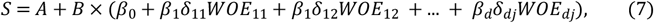

where *β*_*k*_ is the coefficient of feature *k*, and δ_*kj*_ indicates whether the observation is within category *j* of variable *k*. We round the score of each category to the nearest integer, that is:

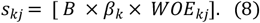

Then, the risk score for a patient with variable vector (*x*_1_, *x*_2_, …, *x*_*d*_) is given by:

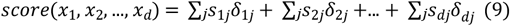

To calculate *score*(*x*_1_, *x*_2_, …, *x*_*d*_), we first determine the category of each model variable *x*_*k*_, and then use the score of that category. Subsequently, the conversion of the risk score to hospitalization probability is through the following logit function:

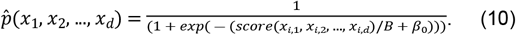

The values of parameters A and B are determined based on two inputs: (i) the target score *S*_0_ corresponding to a certain odds ratio *θ*_0_ for the outcome (i.e., unplanned UTI hospitalization), (ii) points to double the odds Δ*S* such that *S*_0_ +Δ*S* corresponds to an odds ratio of 2*θ*_0_. From these two inputs, we have that *S*_0_ = *A* + *B* × *log*(*θ*_0_) and *S*_0_ +Δ*S* = *A* + *B* × *log*(2*θ*_0_). We can solve these equations together to obtain *B* = Δ*S*/*log*(2) and *A* = *S*_0_ ― *log*(*θ*_0_) × Δ*S*/*log*(2). Note that *A* can be set to 0 for multiple choices of *S*_0_ and *θ*_0_ such that *S*_0_ = *log*(*θ*_0_) × *B*.

As can be seen in Eq. (7), the offset parameter A can be used to shift the value of the scores in the positive or negative direction. On the other hand, the scaling parameter *B* controls the range of the scores assigned to each category. A positive *B* ensures that higher scores correspond to higher risk for the outcome. In our analysis for the unplanned UTI hospitalization, we set the points to double the odds Δ*S* as 10, and *A* as 0.

## Results

We select a prediction score threshold that maximizes F10 score because unplanned UTI hospitalizations are rare in our data. The weight assigned to false negatives (i.e., misclassifying patients with an actual unplanned UTI hospitalization) in F10 score is 100 times the weight of false positives (i.e., misclassifying patients without an unplanned UTI hospitalization). This reflects the fact that reviewing the case of a patient predicted as high-risk for unplanned UTI hospitalization is much more cost-effective than overlooking it and having to deal with a potential hospitalization in the future.

Fig 1 shows the F10 score for each method obtained by using different prediction thresholds over the training set. If the threshold is too low, the model predicts almost all the patients as high risk for hospitalization. In contrast, a large threshold would result in predicting no hospitalization for most of the patients.

**Fig 1.**
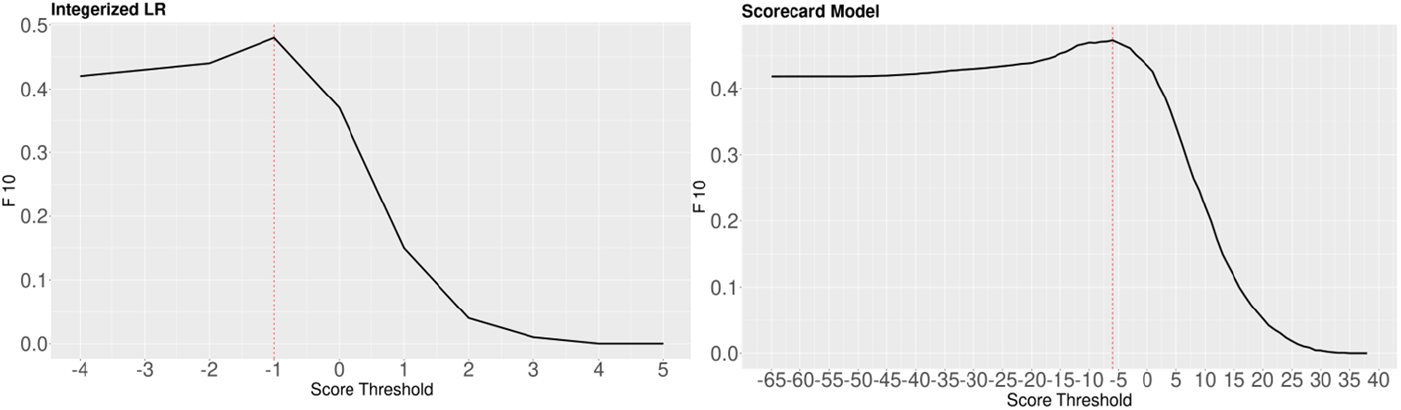
The F10 score for different prediction score thresholds. Dashed line shows the total score where the maximum F10 over the training set is obtained.

As seen in Fig 1, if a lower threshold is chosen, the overall F10 score is fairly robust. Using the thresholds obtained from the F10 analysis, the overall performance of the integerized LR and credit scorecard methods on the test data are summarized in Table 3.

**Table 3.** Prediction performance of the risk score models and the logistic regression (LR) model.

|  | AUC | Recall | Precision | F1 Score | F10 Score |
| --- | --- | --- | --- | --- | --- |
| **LR** | 0.663 | 0.918 | 0.009 | 0.018 | 0.457 |
| **Integerized LR** | 0.67 | 0.903 | 0.009 | 0.019 | 0.467 |
| **Scorecard Model** | 0.688 | 0.855 | 0.01 | 0.02 | 0.467 |

Table 4 presents the scores of the variables selected by the Integerized LR model. A higher score is associated with a higher risk of hospitalization.

**Table 4.** Risk scores of the variables in the Integerized LR method.

| Attribute Name | Score |
| --- | --- |
| Delirium, dementia, amnestic and other cognitive disorders | 1 |
| CCU within last 3 months | 2 |
| ICU within last 3 months | 2 |
| No inpatient comorbidities in the past month | -3 |
| No SNF comorbidities in the past month | -1 |
| Less than \$30 total inpatient and SNF costs in the past 6 months | 1 |

Table 5 presents the scores of the variables selected by the credit scorecard model. Note that there is a score assigned to each category within a variable because this model treats different categories as related rather than separate variables. Furthermore, categories are created in such a way that their scores are monotonic.

**Table 5.** Risk scores of the variables in the credit scorecard model (the cost values are rounded for brevity)

| age | score | Elixhauser inpatient score | score | Elixhauser SNF score | score | Carrier cost within last 6 months | score | Total inpatient and outpatient cost within last 6 months | score |
| --- | --- | --- | --- | --- | --- | --- | --- | --- | --- |
| < 65 | -5 | 0 | -29 | 0 | -2 | <30 | 6 | <30 | 7 |
| 65 - 69 | -4 | 1-2 | -2 | 1 | 1 | [30-600) | 3 | [30-1800) | 4 |
| 70 - 74 | -3 | 3 | -1 | 2 | 2 | [600, 1200) | 2 | [1800-3600) | -1 |
| 75 - 79 | -1 | 4-6 | 2 | 3-4 | 3 | [1200-1300) | 0 | [3600-82500) | -3 |
| 80 - 84 | 0 | >6 | 3 | 5-7 | 4 | >=1300 | -2 | [82500 – 187900) | -10 |
| > 84 | 2 |  |  | >7 | 7 |  |  | >=187900 | -18 |

| Delirium, dementia, amnesic and other cognitive disorders | score | UTI infection diagnosis more than 6 months | score | Bacterial infection diagnosis more than 6 months | score | Recent provider's rate of readmission within 30 days after discharge from hospital | score | ICU within last 3 months | score |
| --- | --- | --- | --- | --- | --- | --- | --- | --- | --- |
| No | -3 | No | -2 | No | -1 | <12.9 | -3 | No | -1 |
| Yes | 4 | Yes | 1 | Yes | 4 | [12.9 - 14.7) | 0 | Yes | 21 |
|  |  |  |  |  |  | [14.7-17.5) | 1 |  |  |
|  |  |  |  |  |  | >=17.5 | 6 |  |  |

Based on the selected modeling parameters, the range of the total score for the integerized LR model is [-4,6] while the range for the credit scorecard is [-65, 40]. These models can be used to calculate a total risk score for a patient and make a classification for unplanned UTI hospitalization (0 or 1) based on a threshold value. We illustrate their usage in Appendix Tables A1 and A2.

To assess how well the proposed scoring methods differentiate between the patients with and without unplanned UTI hospitalization, we examine Fig 2, which illustrates the score distribution within each group for both methods. The cyan and red bars represent patients with and without UTI hospitalization, respectively. The figure illustrates some aspects that relate to AUC and recall. Since integerized LR has higher recall in Table 3, it has more true hospitalizations above the threshold. On the other hand, the scorecard model has better AUC. In Fig 2, this is related to the overlap between the cyan and red bars. The scoring method that exhibits less overlap between these bars performs better in distinguishing the risk of patients with and without unplanned UTI hospitalization. We quantify the overlap between the density plots of scores for patients with and without hospitalization based on the ratio of the overlap area to the total area under each plot.

**Fig 2.**
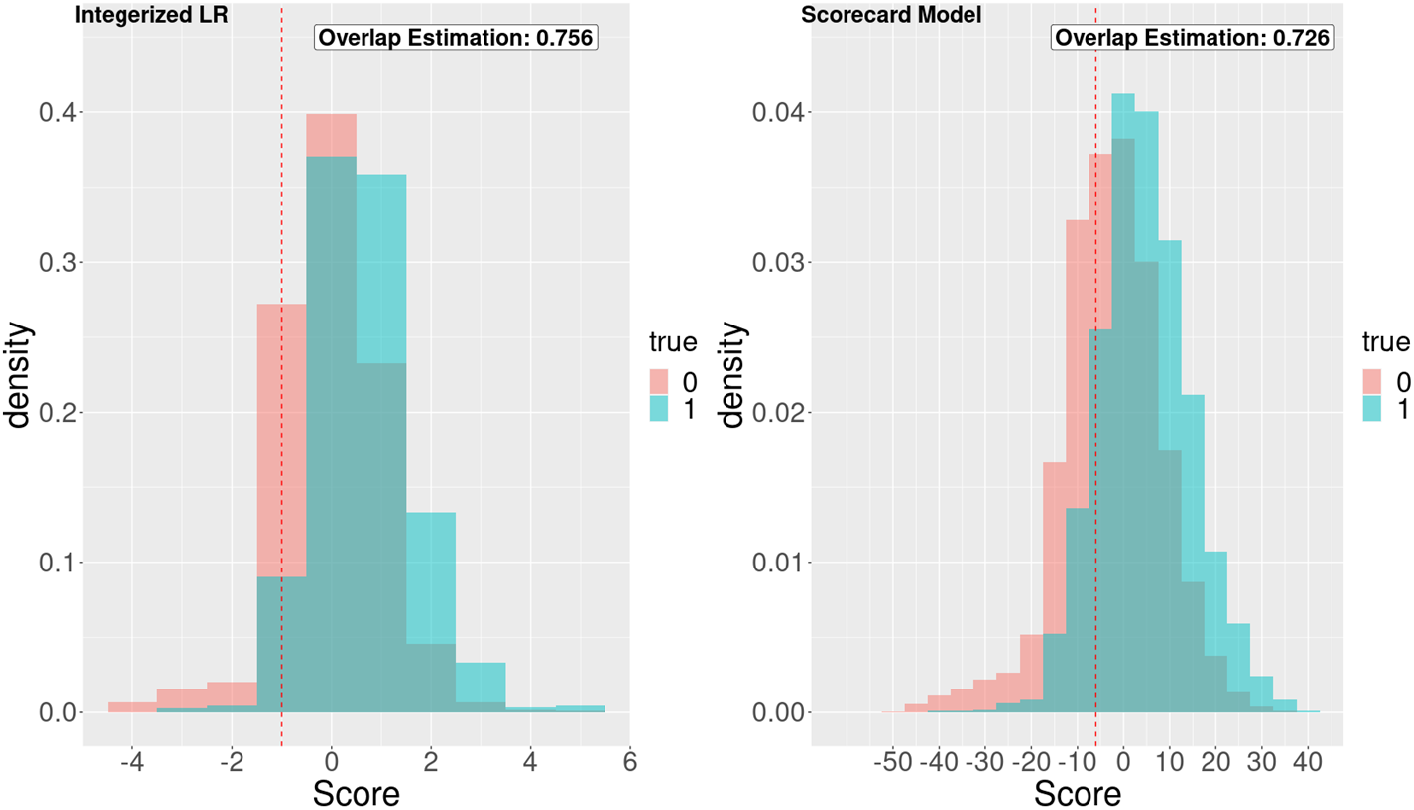
Distribution of the scores for each method. The cyan and red bars represent patients with and without UTI hospitalization, respectively.

Policymakers can leverage risk score models for individualized interventions, such as phone calls [46], and preventive care programs [47], to reduce the burden of unplanned UTI hospitalizations. Population average prescriptive effect (PAPE) and area under the prescriptive effect curve (AUPEC) are two metrics that evaluate the efficiency of individualized treatment rules (ITRs) in comparison to random allocation of treatments [48]. Inspired by these metrics, we randomly select a proportion of patients (e.g., 0 to 100%) from the overall data set to allocate an intervention. We repeat this sampling process 100 times and calculate the recall in each replication. For the allocation of intervention based on the risk score, we select the individuals with the highest score to receive the intervention and calculate the recall. That is, the intervention is either allocated to a randomly selected subpopulation or allocated to the same number of individuals with the highest risk scores to show the benefit of using the risk score. Fig 3 illustrates the results of this experiment. The area between the random selection and the credit scorecard model is 0.187, and it is 0.179 for integerized LR.

**Fig 3.**
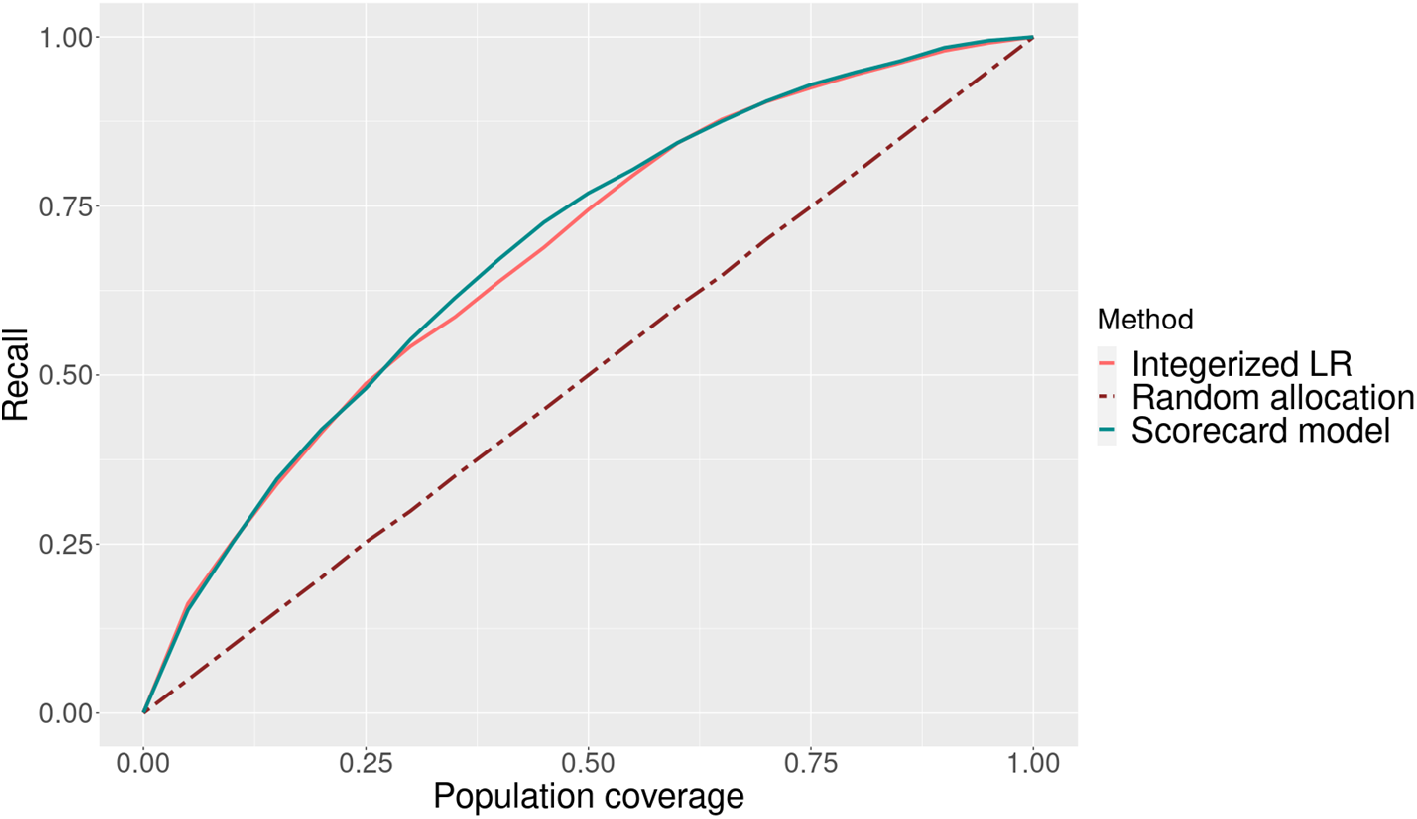
Improvement in recall from using each risk score model to allocate an intervention compared to random allocation.

## Discussion

We proposed two practical risk score models for evaluating the risk of unplanned UTI hospitalization. Both models utilize logistic regression, but they pre-process variables and allocate scores in different ways. We also compared the approach of prioritizing interventions based on predicted risk scores to that of not prioritizing (i.e., random allocation).

Both risk score models demonstrate relatively strong prediction performance given the use of administrative claims data on a population where unplanned UTI hospitalizations are rare (i.e., 0.71%). The precision in Table 3 is low due to the imbalanced nature of our dataset. Overall, our prediction performance results are similar to the ones reported in the literature using claims data from CMS. Specifically, Mao et al. [19] reported 0.63 AUC for the same outcome using a similar CMS dataset which contains beneficiaries both with and without past UTI diagnosis. They improved this result to 0.72 by clustering beneficiaries and training a model for each cluster. The prediction performance results are also comparable to the performance results for the prediction of unplanned pneumonia hospitalization using a similar data set from CMS [49]. The integerized LR model performs better on recall while the credit scorecard performs better on AUC. It is possible that the specific choice of prediction thresholds relates to this trade-off. In Fig 2, the performance of the credit scorecard model with respect to overlap is slightly better than the integerized LR. This may be due to the differences in the range of score values. As seen in Fig 3, both scoring models offer a significant improvement in the percentage of true cases covered when allocating an intervention based on predicted risk, as compared to random allocation.

The proposed risk score models select several common variables, including diagnosis of delirium, dementia, and other cognitive disorders, admission to ICU in the past 3 months, and SNF and inpatient elixhauser scores. Studies in the literature show that variables such as age, history of the UTI [3], and CCU admissions are associated with higher UTI risk [50]. In the credit scorecard model, the elixhauser inpatient score has the widest range of points assigned. Patients without any comorbidity receive a low score of -29, while patients with more than six comorbidities receive the highest score of 3. This difference indicates that the odds of unplanned UTI hospitalization is estimated to be three times higher for the last category of this variable compared to its first category. For the credit scorecard model, it is worthwhile to note that the variable with the largest positive score (indicating an increased risk of unplanned hospitalization), is ICU admission within the past 3 months with a score of 21. For the integerized LR method, the features with the lowest and highest scores are no inpatient comorbidities in the past month with a low score of -3 and ICU/CCU admission within the past 3 months with a score of 2.

Interestingly, the points assigned in both models show that beneficiaries with lower costs exhibit a higher risk for unplanned UTI hospitalization. One possible explanation for this result is that patients with higher costs are more closely (and recently) monitored, and their medical conditions are managed more effectively. Lu et al. [51] also found that high-cost patients (total outpatient and inpatient costs) with more outpatient visits are at lower risk of potentially preventable hospitalizations and stated that the utilization of outpatient care may reduce hospitalizations through preventive care or more effective disease management. The integerized LR model only considers low costs in the inpatient care to be an important variable, whereas the credit scorecard model also considers the effect of high-cost values when evaluating the risk of unplanned UTI hospitalization.

Overall, the variables selected in the proposed risk score models and the points assigned to them provide valuable insights into the risk of unplanned UTI hospitalization. The interpretability of the variables and the transparency of the score generation ensures that the approach is implementable by design. The specific risk score building method to choose may depend on the preferences of the decision-maker about how variables are treated or scored, and about the approach to determining the range and variability of total scores. For example, the integerized LR method can be used to achieve a minimum total score range (e.g., minimum range of 20). In the credit scorecard method, the focus is on the increase in total score with respect to the increase in the risk of hospitalization. Both are reasonable approaches in practice. There are some limitations of this study. We use a 5% sample of Medicare administrative claims data which doesn’t include laboratory results or vitals. We predict hospitalization risk at a monthly level, and only conduct analysis for individuals with sufficient Medicare data available.

## Conclusion

We introduce two methods to build risk score models for unplanned UTI hospitalization utilizing claims data from Medicare limited data sets containing demographics, clinical history, and health care utilization information. We augment this administrative claims data by community-level variables and provider quality metrics obtained from publicly available data sources. We focus on patients with a history of UTI diagnosis. By implementing the integerized LR and credit scorecard models, we assign a risk score to up to 10 important variables associated with unplanned UTI hospitalization. Our findings emphasize the effectiveness of risk score models as practical tools for identifying high-risk patients and provide a quantitative assessment of the significance of various risk factors in unplanned UTI hospitalizations such as admission to ICU in the last 3 months, cognitive disorders and low inpatient, outpatient and carrier costs in the last 6 months.

In future studies, accounting for temporal changes in patients’ conditions and risk factors may lead to improved prediction results. Additionally, addressing the imbalance in the dataset requires exploring advanced techniques like ensemble learning or neural networks [52,53] as traditional approaches such as over sampling and under sampling did not show improved performance in our experiments. Moreover, we presented the results only for a subset of the patients who had a past diagnosis of UTI. The proposed methods can be implemented for other patient groups. Then, the base-level risk of different patient groups can be compared by including the intercept in the total risk score calculation.

## Data Availability

The data underlying the results presented in the study are available from Centers for Medicare and Medicaid Services (data.cms.gov).

## Acknowledgments

The authors thank the Centers for Medicare and Medicaid Services (CMS) for making available the medical claims data utilized in this study.

## Appendix

**Table A 1.** Integerized LR scoring table shows the method obtains the correct prediction outcome for these two patients.

| Criteria | Score | Pos. Patient | Neg. Patient |
| --- | --- | --- | --- |
| Was the patient diagnosed with cognitive disorders, such as dementia, delirium? | 1 | 0 | 0 |
| Was the patient admitted to CCU in the past three months? | 2 | 0 | 0 |
| Was the patient admitted to ICU in the past three months? | 2 | 0 | 0 |
| Did the patient have low inpatient comorbidities in the past month? | -3 | 0 | 0 |
| Did the patient have low SNF comorbidities in the past month? | -1 | -1 | -1 |
| Was the total inpatient and SNF costs of the patient in the past six | 1 | 1 | 0 |
| months low? |  |  |  |
| **Total Score (threshold = -1, where score that is greater predicts event)** |  | 0 | -1 |
| **Prediction** |  | 1 | 0 |

**Table A 2.** Credit Scorecard scoring table shows the method obtains the correct prediction outcome for these two patients)

| Criteria | Pos. Patient category | Pos. Patient Score | Neg. Patient category | Neg. Patient score |
| --- | --- | --- | --- | --- |
| Age | 4 | -1 | 5 | 0 |
| Elixhauser inpatient score | 2 | -2 | 2 | -2 |
| Elixhauser SNF score | 1 | -2 | 1 | -2 |
| Delirium, dementia, amnesic and other cognitive disorders | 0 | -3 | 0 | -3 |
| UTI infection diagnosis more than 6 months | 1 | 1 | 1 | 1 |
| Bacterial infection diagnosis more than 6 months | 0 | -1 | 0 | -1 |
| ICU within last 3 months | 0 | -1 | 0 | -1 |
| Rate of readmission within 30 days after discharge from hospital | 3 | 1 | 3 | 1 |
| Carrier cost within 6 months | 3 | 2 | 5 | -2 |
| Total inpatient and outpatient cost during last 6 months | 1 | 7 | 4 | -3 |
| **Total Score (threshold = -6, where score that is greater predicts event)** |  | 1 |  | -12 |
| **Prediction** |  | 1 |  | 0 |

## Reference

1. McLellan LK, Hunstad DA. Urinary Tract Infection: Pathogenesis and Outlook. Trends Mol Med. 2016;22: 946. doi:10.1016/J.MOLMED.2016.09.003

2. Flores-Mireles A, Hreha TN, Hunstad DA. Pathophysiology, Treatment, and Prevention of Catheter-Associated Urinary Tract Infection. Top Spinal Cord Inj Rehabil. 2019;25: 228–240. doi:10.1310/SCI2503-228

3. Flores-Mireles AL, Walker JN, Caparon M, Hultgren SJ. Urinary tract infections: epidemiology, mechanisms of infection and treatment options. Nat Rev Microbiol. 2015;13: 269–284. doi:10.1038/NRMICRO3432

4. Öztürk R, Murt A. Epidemiology of urological infections: a global burden. World J Urol. 2020;38: 2669–2679. doi:10.1007/S00345-019-03071-4/TABLES/2

5. Holm A, Cordoba G, Aabenhus R. Prescription of antibiotics for urinary tract infection in general practice in Denmark. Scand J Prim Health Care. 2019;37: 83. doi:10.1080/02813432.2019.1569425

6. Unroe KT, Carnahan JL, Hickman SE, Sachs GA, Hass Z, Arling G. The Complexity of Determining Whether a Nursing Home Transfer Is Avoidable at Time of Transfer. J Am Geriatr Soc. 2018;66: 895–901. doi:10.1111/JGS.15286

7. Will JC, Valderrama AL, Yoon PW. Preventable hospitalizations for congestive heart failure: Establishing a baseline to monitor trends and disparities. Prev Chronic Dis. 2012;9. doi:10.5888/PCD9.110260

8. Kimberly W. McDermott, H. Joanna Jiang. Characteristics and Costs of Potentially Preventable Inpatient Stays, 2017. Available: https://hcup-us.ahrq.gov/reports/statbriefs/sb259-Potentially-Preventable-Hospitalizations-2017.pdf

9. Medina M, Castillo-Pino E. An introduction to the epidemiology and burden of urinary tract infections. Ther Adv Urol. 2019;11: 1756287219832172. doi:10.1177/1756287219832172

10. Hooton TM. Recurrent urinary tract infection in women. Int J Antimicrob Agents. 2001;17: 259–268. doi:10.1016/S0924-8579(00)00350-2

11. Tandogdu Z, Cai T, Koves B, Wagenlehner F, Bjerklund-Johansen TE. Urinary Tract Infections in Immunocompromised Patients with Diabetes, Chronic Kidney Disease, and Kidney Transplant. Eur Urol Focus. 2016;2: 394–399. doi:10.1016/J.EUF.2016.08.006

12. Szweda H, Jóźwik M. Urinary tract infections during pregnancy - an updated overview. Dev Period Med. 2016;20: 263–272.

13. Caljouw MAA, den Elzen WPJ, Cools HJM, Gussekloo J. Predictive factors of urinary tract infections among the oldest old in the general population. A population-based prospective follow-up study. BMC Med. 2011;9: 1–8. doi:10.1186/1741-7015-9-57/FIGURES/1

14. Bono MJ, Leslie SW, Reygaert WC. Urinary Tract Infection. StatPearls. 2022 [cited 14 Jun 2023]. Available: https://www.ncbi.nlm.nih.gov/books/NBK470195/

15. Taylor RA, Moore CL, Cheung KH, Brandt C. Predicting urinary tract infections in the emergency department with machine learning. PLoS One. 2018;13: e0194085. doi:10.1371/JOURNAL.PONE.0194085

16. Møller JK, Sørensen M, Hardahl C. Prediction of risk of acquiring urinary tract infection during hospital stay based on machine-learning: A retrospective cohort study. PLoS One. 2021;16: e0248636. doi:10.1371/JOURNAL.PONE.0248636

17. Mancini A, Vito L, Marcelli E, Piangerelli M, De Leone R, Pucciarelli S, et al. Machine learning models predicting multidrug resistant urinary tract infections using “dsaaS.” BMC Bioinformatics. 2020;21: 1–12. doi:10.1186/S12859-020-03566-7/TABLES/3

18. Gopichand P, Agarwal G, Natarajan M, Mandal J, Deepanjali S, Parameswaran S, et al. In vitro effect of fosfomycin on multi-drug resistant gram-negative bacteria causing urinary tract infections. Infect Drug Resist. 2019;12: 2005–2013. doi:10.2147/IDR.S207569

19. Mao L, Vahdat K, Shashaani S, Swann JL. Personalized Predictions for Unplanned Urinary Tract Infection Hospitalizations with Hierarchical Clustering. Springer Proceedings in Business and Economics. 2022; 453–465. doi:10.1007/978-3-030-75166-1_34/TABLES/3

20. Rudin C, Chen C, Chen Z, Huang H, Semenova L, Zhong C. Interpretable Machine Learning: Fundamental Principles and 10 Grand Challenges. Stat Surv. 2021;16: 1–85. doi:10.1214/21-SS133

21. Adadi A, Berrada M. Explainable AI for Healthcare: From Black Box to Interpretable Models. Advances in Intelligent Systems and Computing. 2020;1076: 327–337. doi:10.1007/978-981-15-0947-6_31/FIGURES/1

22. Luo Y, Tseng H-H, Cui S, Wei L, Haken RK Ten, Naqa I El. Balancing accuracy and interpretability of machine learning approaches for radiation treatment outcomes modeling. BJR Open. 2019;1. doi:10.1259/BJRO.20190021

23. Han JT, Park IS, Kang SB, Seo BG. Developing the High-Risk Drinking Scorecard Model in Korea. Osong Public Health Res Perspect. 2018;9: 231. doi:10.24171/J.PHRP.2018.9.5.04

24. Than M, Flaws D, Sanders S, Doust J, Glasziou P, Kline J, et al. Development and validation of the emergency department assessment of chest pain score and 2h accelerated diagnostic protocol. EMA - Emergency Medicine Australasia. 2014;26: 34–44. doi:10.1111/1742-6723.12164/SUPPINFO

25. Six AJ, Backus BE, Kelder JC. Chest pain in the emergency room: value of the HEART score. Netherlands Heart Journal. 2008;16: 191. doi:10.1007/BF03086144

26. Markov A, Seleznyova Z, Lapshin V. Credit scoring methods: Latest trends and points to consider. The Journal of Finance and Data Science. 2022;8: 180–201. doi:10.1016/J.JFDS.2022.07.002

27. Zeng J, Ustun B, A CR-J of the RSSocietyS, 2017 undefined. Interpretable classification models for recidivism prediction. JSTOR. [cited 18 Jun 2023]. Available: https://www.jstor.org/stable/44682879

28. Indicators A. Patient Safety Indicators Technical Specifications. 2007 [cited 14 Jun 2023]. Available: https://qualityindicators.ahrq.gov/downloads/modules/psi/v43/techspecs/psi%20appendices.pdf

29. Centers for Medicare & Medicaid Services. Announcement of Requirements and Registration for “Artificial Intelligence Health Outcomes Challenge.” [cited 18 Jun 2023]. Available: https://innovation.cms.gov/files/x/aichallenge-pubnotice_08_13_2020.pdf

30. Census Bureau Data. [cited 14 Jun 2023]. Available: https://data.census.gov/

31. CDC - BRFSS. [cited 14 Jun 2023]. Available: https://www.cdc.gov/brfss/index.html

32. County Health Rankings & Roadmaps. [cited 14 Jun 2023]. Available: https://www.countyhealthrankings.org/

33. Provider Data Catalog. [cited 14 Jun 2023]. Available: https://data.cms.gov/provider-data/?redirect=true

34. Search | Provider Data Catalog. [cited 14 Jun 2023]. Available: https://data.cms.gov/provider-data/search?theme=Nursing%20homes%20including%20rehab%20services

35. Weekly U.S. Influenza Surveillance Report | CDC. [cited 14 Jun 2023]. Available: https://www.cdc.gov/flu/weekly/index.htm

36. Shams SF, Eidgahi ES, Lotfi Z, Khaledi A, Shakeri S, Sheikhi M, et al. Urinary tract infections in kidney transplant recipients 1st year after transplantation. J Res Med Sci. 2017;22. doi:10.4103/1735-1995.200274

37. Will JC, Nwaise IA, Schieb L, Zhong Y. Geographic and racial patterns of preventable hospitalizations for hypertension: Medicare beneficiaries, 2004-2009. Public Health Rep. 2014;129: 8–18. doi:10.1177/003335491412900104

38. Moghadamyeghaneh Z, Stamos MJ, Stewart L. Patient Co-Morbidity and Functional Status Influence the Occurrence of Hospital Acquired Conditions More Strongly than Hospital Factors. Journal of Gastrointestinal Surgery. 2019;23: 163–172. doi:10.1007/S11605-018-3957-9/FIGURES/2

39. Chan JK, Gardner AB, Mann AK, Kapp DS. Hospital-acquired conditions after surgery for gynecologic cancer — An analysis of 82,304 patients. Gynecol Oncol. 2018;150: 515–520. doi:10.1016/J.YGYNO.2018.07.009

40. Carter MW. Factors Associated with Ambulatory Care—Sensitive Hospitalizations among Nursing Home Residents. http://dx.doi.org/101177/0898264303015002001. 2003;15: 295–331. doi:10.1177/0898264303015002001

41. Saver BG, Wang CY, Dobie SA, Green PK, Baldwin LM. The central role of comorbidity in predicting ambulatory care sensitive hospitalizations*. Eur J Public Health. 2014;24: 66–72. doi:10.1093/EURPUB/CKT019

42. Regression Shrinkage and Selection via the Lasso on JSTOR. [cited 15 Jun 2023]. Available: https://www.jstor.org/stable/2346178

43. Siddiqi N. Credit risk scorecards: developing and implementing intelligent credit scoring. 2012. Available: https://books.google.com/books?hl=en&lr=&id=SEbCeN3-kEUC&oi=fnd&pg=PT7&dq=Credit+Risk+Scorecards:+Developing+and+Implementing+Intelligent+Credit+Scoring+&ots=RvWS3OcQkT&sig=oEhGkzaTFY5rr4UvulWI4eR3aiI

44. Lee CY, Koh SK, Lee MC, Pan WY. Application of Machine Learning in Credit Risk Scorecard. Communications in Computer and Information Science. 2021;1489 CCIS: 395–410. doi:10.1007/978-981-16-7334-4_29/TABLES/4

45. Rabobank PM, Rabobank VT. Monotone optimal binning algorithm for credit risk modeling. 2017 [cited 15 Jun 2023]. doi:10.13140/RG.2.2.31885.44003

46. Parmar P, Mackie D, Varghese S, Cooper C. Use of Telemedicine Technologies in the Management of Infectious Diseases: A Review. Clinical Infectious Diseases. 2015;60: 1084–1094. doi:10.1093/CID/CIU1143

47. Infection Prevention and Control - Physiopedia. [cited 14 Jun 2023]. Available: https://www.physio-pedia.com/Infection_Prevention_and_Control

48. Imai K, Li ML. Experimental Evaluation of Individualized Treatment Rules. J Am Stat Assoc. 2021;2023: 242–256. doi:10.1080/01621459.2021.1923511/SUPPL_FILE/UASA_A_1923511_SM6049.PDF

49. Data Analytics and Decision Making: Evaluating Risk and Burden Associated with Infectious Respiratory Diseases. [cited 17 Jun 2023]. Available: https://repository.lib.ncsu.edu/handle/1840.20/39958

50. Parida S, Mishra SK. Urinary tract infections in the critical care unit: A brief review. Indian J Crit Care Med. 2013;17: 370. doi:10.4103/0972-5229.123451

51. Lu S, Zhang Y, Zhang L, Klazinga NS, Kringos DS. Characterizing Potentially Preventable Hospitalizations of High-Cost Patients in Rural China. Front Public Health. 2022;10: 804734. doi:10.3389/FPUBH.2022.804734/BIBTEX

52. Sun B, Chen H, Wang J, Xie H. Evolutionary under-sampling based bagging ensemble method for imbalanced data classification. Front Comput Sci. 2018;12: 331–350. doi:10.1007/S11704-016-5306-Z/METRICS

53. Pattanayak D, Patel K. Generative Adversarial Networks: Solution for Handling Imbalanced Datasets in Computer Vision. 2022 International Conference for Advancement in Technology, ICONAT 2022. 2022. doi:10.1109/ICONAT53423.2022.9725995

